# An extract of hops (*Humulus lupulus* L.) modulates gut peptide hormone secretion and reduces energy intake in healthy weight men: a randomised, cross-over clinical trial

**DOI:** 10.1101/2021.06.25.21259514

**Authors:** Edward G Walker, Kim R Lo, Malcolm C Pahl, Hyun Sang Shin, Claudia Lang, Mark W Wohlers, Sally D Poppitt, Kevin H Sutton, John R Ingram

## Abstract

**Background:** Gastrointestinal enteroendocrine cells express a range of chemosensory receptors involved in detecting the chemical composition of food during digestion. These receptors, including bitter taste receptors (T2Rs), may play an important role in regulating gut function and appetite.

**Objective:** To establish the ability of Amarasate^®^, a bitter supercritical CO_2_ extract of hops (*Humulus lupulus* L.) to modify acute energy intake, appetite and hormonal responses and establish a site of action.

**Design:** Nineteen healthy-weight (BMI = 23.5 ± 0.3 kg/m^2^) male volunteers completed a randomised three-treatment, double blind, cross-over study with a 1 week washout between treatments. Overnight-fasted participants were cannulated and provided with a standardised 2 MJ breakfast meal at 0900h. Treatments comprised a vehicle control (Placebo) or 500 mg of hops extract administered in either delayed release capsules (Duodenal) at 1100 h or quick release capsules (Gastric) at 1130 h. *Ad libitum* energy intake was recorded at an outcome meal (1200 h) and afternoon snack (1400 h), with blood samples taken and subjective ratings of appetite, gastrointestinal discomfort, vitality, meal palatability and mood assessed throughout the day.

**Results:** Compared with placebo, both gastric and duodenal treatments significantly reduced (p < 0.05) total *ad libitum* energy intake by 911 ± 308 kJ and 944 ± 309 kJ, respectively. Both gastric and duodenal treatments significantly increased (p < 0.05) pre-meal ghrelin and post-prandial CCK, GLP-1 and PYY responses while reducing postprandial insulin, GIP and PP secretion with no significant impact on glycemia. In addition, gastric and duodenal treatments produced small but significant (p < 0.05) changes in vitality and gastrointestinal discomfort (e.g. nausea, bloating, abdominal discomfort) with mild-moderate adverse GI symptoms reported in the gastric treatment only. However, no significant treatment effects were observed for any subjective measures of appetite or meal palatability.

**Conclusion:** Both gastric and duodenal delivery of Amarasate^®^ modulate the release of hormones involved in appetite and glycaemic regulation, providing a potential “bitter brake” on energy intake in healthy-weight men.

## INTRODUCTION

Control of energy intake (EI) is central to the success of interventions designed to manage body weight (1) and the consequences of obesity (2-6). The gastrointestinal (GI) tract expresses an array of chemosensory receptors and transporters that provide critical inputs into the acute regulation of energy intake, detecting and relaying to the brain the location, chemical composition and concentration of nutritive and non-nutritive compounds in the gut (7, 8). Obesity and poor weight loss outcomes are associated with impaired gut-brain axis signalling (9-13) which may contribute to overeating and poor adherence to dietary restriction (14-17). Approaches that restore or enhance gut-brain axis signalling may address this underlying feedback dysregulation. Indeed, enhancement of gut-brain axis signalling may explain many of the benefits of gastric bypass surgery (18), dietary strategies (e.g. high fibre/protein) and pharmaceutical interventions (19, 20) on the control of EI. Importantly, GI chemosensory mechanisms are readily accessible to dietary manipulation and represent an unexploited source of weight management targets (21-23).

Bitter taste receptors (T2R) comprise a family of 25 G protein-coupled receptors that are expressed in multiple tissues, including enteroendocrine cells (EEC) of the GI tract (24-26) and are thought to have evolved a chemosensory role in the detection of potential harmful substances, limiting their ingestion and absorption (27, 28). *In vitro*, T2R agonists stimulate the release of peptide hormones, such as ghrelin, cholecystokinin (CCK) and glucagon-like peptide-1 (GLP-1), from gut enteroendocrine cells (29-32). These gut peptide hormones play a key role the homeostatic regulation of appetite, energy intake, gut function, hedonic food perceptions and nutient metabolism (33-37). A number of clinical studies using either encapsulation or intragastric and intraduodenal infusion of bitter tastents have demonstrated effects ranging from increased gut peptide secretion, reduced energy intake or rate of gastric emptying, modifications in subjective ratings of hunger and fullness, and altered glycemic regulation (38-43), although these anorexigeneic effects are inconsistent (43-46), necessitating further investigation of this response.

Hops (*Humulus lupulus L*.) contain a range of bitter compounds including α-acids (humulone, adhumulone and cohumulone) and β-acids (lupulone, adlupulone and colupulone) that are known ligands for human bitter taste receptors (47). They have a long history of use as food additives and bittering agents in brewing, as well as in traditional medicine [40, 41] and have been shown *in vitro* to stimulate Ca^2+^-dependent CCK release from EEC cells (32).

Administration of hop-derived extracts has also been shown to reduce body weight, fat mass and improve glucose homeostasis in both rodent (48-55) and human studies (39, 50, 56). In addition, our laboratory has demonstrated that administration of a supercritical CO_2_ hop extract can reduce subjective ratings of hunger during water-only fasting (57).

Here we investigate the efficacy and GI site of action of Amarasate^®^, a bitter supercritical CO_2_ extract of hops, to modify acute energy intake, hormonal and glycaemic responses, and subjective ratings of appetite, gastrointestinal discomfort, meal palatability and mood in healthy-weight men.

## METHODS

### Participants

Healthy-weight men (18–55 years old), with a BMI between 20 and 25 kg/m^2^ were recruited by advertisement in the Auckland region, New Zealand. A telephone pre-screening interview to determine eligibility of interested individuals was followed by a screening visit to verify eligibility by measurement of height and weight, assessment of oral bitter taste sensitivity to the hops extract, and determination of health status by self-report and blood tests (HbA1c, liver function, full blood count, iron status).

Participants were excluded if they had a diagnosed medical condition or were on medications known to affect taste, appetite-related parameters, metabolism or gastrointestinal function. Exclusions also applied to participants currently on a weight-loss programme or taking weight-loss medication or who had significant weight loss or gain (>5 kg) within the last six months, were smokers, or had a history of alcohol or drug abuse. Participants with hypersensitivities or allergies to any foods or ingredients included in the study, as well as those that disliked or were unwilling to consume items listed as study foods or were unwilling or unable to comply with the study protocol, or who were participating in another clinical intervention trial, were also excluded.

All participants provided informed consent prior to clinical trial enrolment. Human ethics approval was obtained from the Northern B Health and Disability Ethics committee (ref. 14/NTB/25) and the trial registered at the Australian and New Zealand clinical trials registry (ref. ACTRN12614000434695). The study was conducted at the Consumer and Products Insights facility of The New Zealand Institute for Plant and Food Research Limited (Auckland, New Zealand) in March–June 2014.

### Study design

A randomized, double-blind, placebo-controlled, 3-treatment crossover study design was used to determine the efficacy and GI site of action of Amarasate^®^ to modify acute energy intake, appetite and postprandial hormonal responses in healthy-weight men. The three treatment arms (**Supplemental Table 1**) were: Amarasate^®^ (500 mg) targeted for release into the stomach (gastric), Amarasate^®^ (500 mg) targeted for release in the proximal small intestine (duodenum), and a vehicle control (placebo). Randomisation was conducted using a 3×3 Latin square balanced for treatment order and carryover effects (58, 59).

Three one-day visits were required with a washout period of at least one week between visits. The daily protocol is shown in **Figure 1**. Food intake and subjective measures of appetite, gastrointestinal discomfort, vitality, meal palatability and mood were assessed during fully supervised study days using standard methodology as per the recommendations of Blundell et al. (60).

**Figure 1.**
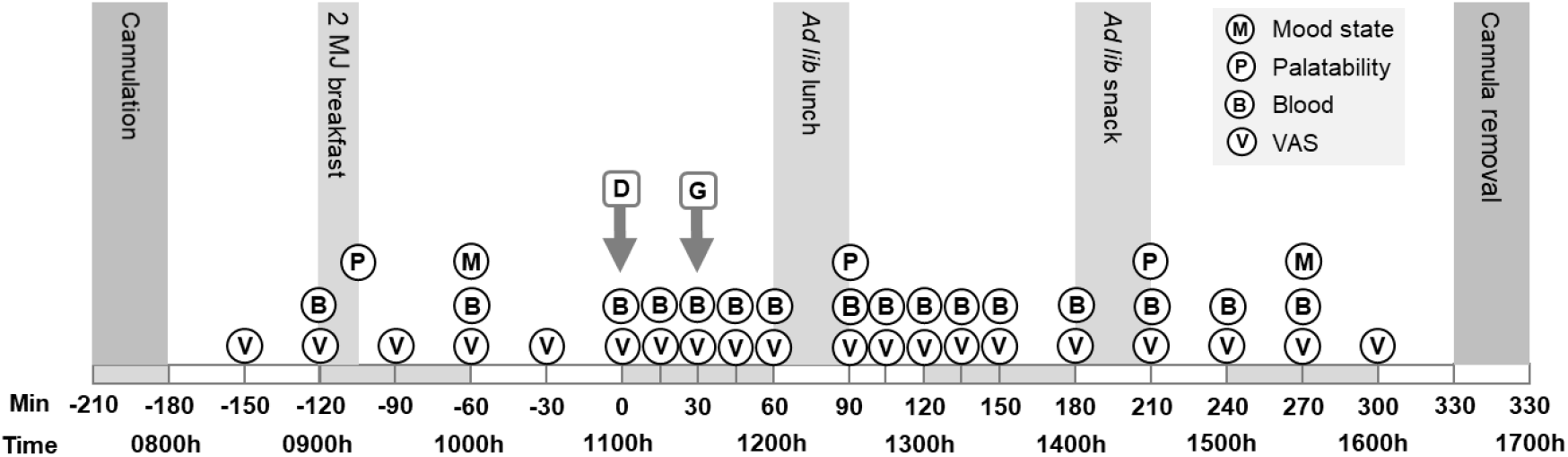
Protocol for study visits 1–3. Participants arrived fasted (0730 h), were cannulated and provided with a fixed energy (2 MJ) breakfast (0900 h) that they had to complete. Treatments along with matched placebo capsules targeting the duodenum (D) or gastric (G) compartments were administered at 1100 h (T=0 min) and 1130 h (T= 30 min), respectively. Participants were provided with a*d libitum* lunch (1200 h) and snack (1400 h) outcome meals and directed to eat until comfortably full. Blood samples (B) and VAS ratings (V) of appetite, thirst, vitality and gastrointestinal discomfort related-measures were collected throughout the day. Ratings of meal palatability (P) were assessed using VAS scales immediately after every meal. Mood state (M) were assessed in the morning and afternoon using the Profile of Mood State questionnaire. VAS, visual analogue scale; *Ad lib, Ad libitum*.

### Treatments

To maintain treatment blinding, all treatments contained two sets of opaque capsules, one set administered at 1100 h were delayed release capsules (DRCaps™, size 0, Capsugel, NJ, USA) designed to release their contents approximately 50–70 min after ingestion, increasing the likelihood of delivery to the duodenum (61). The second set given at 1130 h were standard hydroxypropyl-methylcellulose capsules (Vcaps™, size 0, Capsugel) designed to release in the stomach. The timing of capsule administration was chosen so that the treatment capsules would probably have released their contents in the stomach or duodenum before the *ad libitum* lunch (1200 h).

The three treatment groups were as follows: **Placebo** – two vehicle control delayed-release capsules (1100 h) followed by two vehicle control standard-release capsules (1130 h); **Gastric** – two vehicle control delayed-release capsules (1100 h) followed by two Amarasate^®^ standard-release capsules (1130 h); **Duodenum** – two Amarasate^®^ delayed-release capsules (1100 h) followed by two vehicle control standard-release capsules (1130 h).

Each Amarasate^®^ treatment comprised two capsules, each containing 250 mg of a commercially available food-safe supercritical CO_2_ extract of hop cones (*Humulus lupulus* L. ‘Pacific Gem’) sourced from New Zealand Hops Ltd, NZ, mixed with 125 mg of canola oil as an excipient (a 2:1 hops:oil ratio). The vehicle control capsules utilised in the placebo treatment and for blinding in the gastric and duodenal treatments contained 125 mg of canola oil. All capsules were filled in-house using the Capsugel Profiller system (Capsugel, Morristown New Jersey, USA) with a coefficient of variation of 2% for loading accuracy.

The α- and β-acid composition of the hop supercritical CO_2_ extract comprised 51.5% total α-acids (cohumulone 21.1%, humulone 22.3% and adhumulone 8.2%), and 28.3% total β-acids (colupulone 19.7%, lupulone 6.0% and adlupulone 3.1%) as determined by high performance liquid chromatography (HPLC) (**Supplemental Figure 1**) with reference to the American Brewing Association ICE-3 standard as described in (62). The α- and β-acid composition of the Amarasate^®^ formulation has previously been shown to remain stable over the duration of use in the current study (57).

### Study visits 1–3

Participants arrived at the study facility by 0730 h on test days in an overnight fasted state (no food or drinks apart from water since 2200 h) having abstained from excessive exercise or alcohol consumption the day before. An indwelling venous cannula was inserted into a forearm vein for repeated blood collection. **Figure 1** shows the study visit protocol including timing of meals, treatment administration, and the collection of blood and behavioural measures. During free time between the meals and questionnaires, participants remained inside the facility but were free to read, watch TV or access the internet on their own devices. Participants were free to leave the facility after completion of the final study questionnaire and removal of the cannula at 1600 h.

### Fixed energy breakfast, ad libitum meals, and EI

The fixed energy (2 MJ) breakfast (**Supplemental Table 2**) consisted of puffed rice cereal with low fat milk and white bread with margarine and jam. Participants were instructed to consume the entire breakfast within 15 min (verified by visual inspection). The outcome *ad libitum* lunch (1200 h, T= 60 min) was a savoury buffet restricted to a beef and tomato pasta sauce and boiled pasta spirals with water (250 mL). Ham sandwiches, cut into quarters with the crusts removed, were provided for the outcome afternoon snack (1400 h, T= 180 min) with water (250 mL). Both *ad libitum* meals were provided in excess, with participants instructed that they had 30 minutes to eat until they were comfortably full. To minimise distractions, all meals were provided in individual booths with participants instructed not to talk, read or use mobile phones or electronic devices and to remain in the booth for the designated time. Meals were weighed by two separate observers before and after consumption and energy, fat, carbohydrate, and protein intake were calculated with the use of the dietary software program FoodWorks (Professional Edition, version 5; Xyris Software). All meals were designed to have low phytochemical content to minimise non-specific effects on appetite (63).

### Behavioural measures

Visual analogue scales (VAS) were used to assess subjective feelings of hunger, fullness, satiety, and prospective consumption following the methodology outlined in Blundell et al. (60, 64). Additional VAS were used to assess thirst; measures of vitality (energy levels and relaxation); GI discomfort including nausea, urge to vomit, bloating, abdominal discomfort, and heartburn (adapted from (65)); and meal palatability (64) (pleasantness, visual appeal, smell, taste, aftertaste and overall palatability). The VAS questions and anchor statements are provided in **Supplemental Table 3**. Participants marked their responses by placing a vertical line across the 100-mm scale according to subjective feelings, with responses recorded to the nearest mm.

Changes in mood states were assessed at 1000 h (T = -60) and 1530 h (T = 270) using the original version of the Profile of Mood States (POMS) questionnaire (66), a 65-item inventory of six subscales: tension-anxiety, depression-dejection, anger-hostility, vigour-activity, fatigue-inertia, and confusion-bewilderment. Participants rated “How are you feeling right now” for each mood descriptor on a 5-point scale anchored by 1 = “not at all” and 5 = “extremely”. The total mood disturbance score was computed by adding the five negative subscale scores (tension, depression, anger, fatigue, confusion) and subtracting the vigour score.

The occurance of adverse symptoms/events were recorded for each study visit with participants describing symptoms and their severity using a three-point scale of mild, moderate or severe. Participants were also asked to recall any delayed symptoms/events during the washout period at their next visit.

### Blood measurements

Blood for peptide hormones analysis was collected into pre-chilled 5-mL EDTA tubes (BD Vacutainer^®^, USA) containing a dipeptidyl-aminopeptidase IV inhibitor (25 µL of a 2 mM solution of Diprotin A, Peptides International, Osaka, Japan) and a general protease inhibitor cocktail (Complete Mini EDTA-free protease inhibitor, Roche, Basel, Switzerland) (182 µL of solution made up of one tablet in 2 mL of water). Blood for plasma glucose analysis was collected into sodium fluoride/potassium oxalate Vacutainer^®^ tubes (BD, USA). Upon collection samples were immediately centrifuged (1500x*g* for 10 min at 4^°^C) and the plasma snap frozen on dry ice before storage at -80^°^C until analysis.

Ghrelin (active), GLP-1 (active), PYY (total), insulin, GIP (total) and pancreatic polypeptide (PP) concentrations were measured using a multiplexed magnetic bead assay (HMHMAG-34K; Merck-Millipore, Massachusetts USA). Samples were assayed in duplicate and plates read using a Magpix™ system (Luminex, USA) with concentrations determined using a 5-parameter curve fit in Analyst 5.1 (Miliplex, USA). Plasma CCK concentrations were determined in duplicate by radioimmunoassay (EURIA-CCK, Eurodiagnostica, Sweden) as per the manufacturer’s instructions, with CCK standards formulated in pooled charcoal stripped human plasma. Assay QC data are given in **Supplementary Table 4**. Plasma glucose was analysed by Lab Services (North Shore Hospital Lab Services, Auckland) using the hexokinase method on a Dimension^®^ Vista 1500 (Siemens AG, Erlangen, Germany).

### Statistical Analysis

A completers only analysis was used to address missing data. Time profile data including VAS ratings and blood biomarkers were analysed with the use of a linear mixed model (SAS software, PROC GLIMMIX function, version 9.4; SAS Institute Inc.) with treatment, sample number, visit number and and treatment order (one of six possible treatment sequences allocated to each subject) and their respective interactions included as fixed effects. Fisher’s protected LSD was used to account for multiple testing. Where there was evidence of a main treatment or treatment x time interaction (p < 0.05), F-tests for treatment differences at each time point were conducted using the ‘slice’ command. Where these were significant (p < 0.05), Fisher’s protected LSD *post hoc* analysis was used for pairwise comparisons between treatments.

Area under the curve (AUC) data were calculated from time 0 to 270 min for blood biomarkers, and from 0 to 300 min for VAS measures, and analysed using a linear mixed model (SAS 9.4) with treatment, visit number and and treatment order as fixed effects.. Where there was evidence of a main treatment effect (p <0.05), Fisher’s protected LSD was used to account for multiple testing of pairwise comparisons between treatments. EI data were analysed in the same way, with models fitted separately for the snack, lunch, and total kJ intake measures. For meal palability measures an additional fixed factor of meal (breakfast, lunch and snack) was included in the linear mixed model, while analysis of POMS subscales included the fixed factor of time (pre/post). Results are presented as means ± SEM; if required, data were log transformed before analysis, with results presented as back-transformed means ± SEM. Statistical significance was assessed at p <0.05.

## Results

### Participants

Of the 20 healthy-weight male participants randomly assigned into the trial, 19 completed all three arms of the study, with one participant excluded for failure to comply with study protocol (see CONSORT flow diagram, **Supplemental Figure 2**). Characteristics of the 19 participants included in the final analysis of energy intake and subjective behavioural measures are shown in **Table 1**. A second participant was excluded from blood sample analysis only because of repeated cannula failures and inability to obtain sufficient blood volume. Hence, data on blood biomarkers is presented for 18 participants.

**Table 1.** Characteristics of the 19 male participants who completed all three treatment arms^1^

| | Mean ± SD | Range |
| --- | --- | --- |
| Age, y | 28.9 ± 10.4 | 18–54 |
| Height, m | 1.80 ± 0.08 | 1.66–1.95 |
| Body weight, kg | 76.1 ± 8.3 | 60.4–94.5 |
| BMI, kg/m<sup>2</sup> | 23.5 ± 1.4 | 20.9–25.0 |
| Ethnicity<sup>2</sup>: |  |  |
| New Zealand European | 13 |  |
| Maori/Pacifica | 2 |  |
| Asian | 3 |  |
| Other | 1 |  |

<sup>1</sup>All measurements were recorded at the screening visit. <sup>2</sup>Ethnicity was assessed by self-report.

### EI at ad libitum meals

The effects of treatment on EI at the outcome *ad libitum* lunch and snack meals are shown in **Figure 2**. Total EI from the two outcome meals showed a highly significant effect of treatment (F_2,34_ = 6.0, p = 0.006), with both the gastric (4320 ± 350 kJ, p= 0.015) and duodenal (4287 ± 350 kJ, p= 0.012) treatments resulting in significant reductions compared with the placebo (5231 ± 350 kJ). A significant effect of treatment (F_2,34_ = 4.0, p = 0.027) was observed at the *ad libitum* snack with a reduction of EI in the duodenal treatment (1423 ± 199 kJ, p=0.044) compared with the placebo (2018 ± 199 kJ), while the values in the gastric treatment (1452 ± 199 kJ, p=0.056) just failed to reach statistical significance. EI at the *ad libitum* lunch showed no significant effect of treatment.

**Figure 2.**
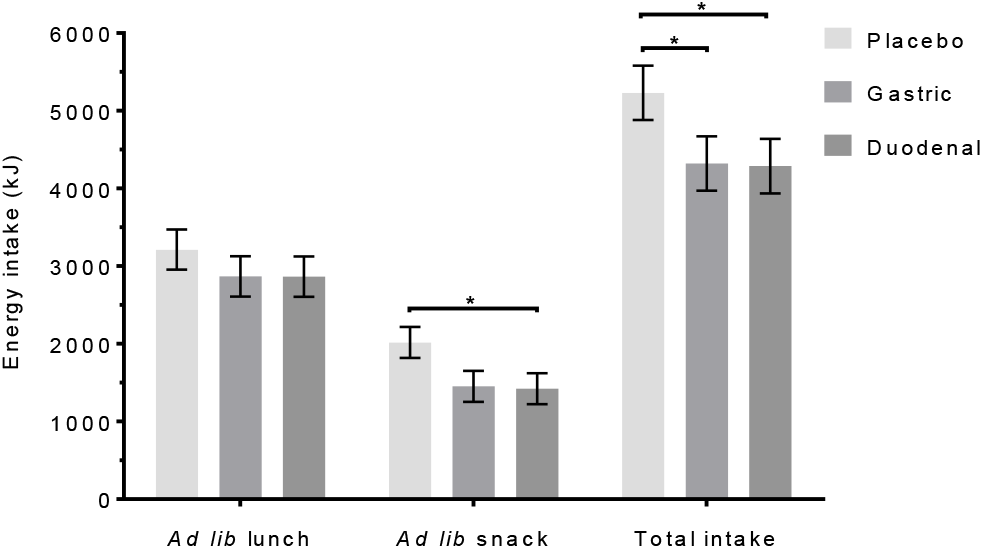
The effect of treatment on *ad libitum* energy intake (kJ) at the outcome lunch (1200 h), snack (1400 h) and the combined intake (Total intake). Treatments comprised either a vehicle control (Placebo) or a formulated hops extract (Amarasate^®^) designed to release in the stomach (Gastric) or in the proximal small intestine (Duodenum). Analysis was conducted using the Mixed procedure (SAS 9.4) with treatment, visit number and treatment order as factors. A significant effect of treatment was observed for both the snack (F_2,34_ = 4.0, *p* = 0.027) and for total intake (F_2,34_= 6.0, *p* = 0.006). Fisher’s LSD *post hoc* pairwise analysis demonstrated a significant (*p* = 0.044) reduction in energy intake for the duodenal treatment compared with the placebo at the snack and for both the gastric (*p* = 0.015) and duodenal (*p* = 0.012) treatments compared with the placebo when assessed as total intake. Values are means ± sem, (n = 19). * p < 0.05.

### Blood parameters

#### Ghrelin, CCK, GLP-1 and PYY

The effects of treatment on plasma concentrations and AUC_0-270 min_ responses of the appetite regulating hormones ghrelin, CCK, PYY and PP are shown in **Figure 3A–D**. All four peptide hormone profiles exhibited predictable changes driven primarily by the timing of meals.

**Figure 3.**
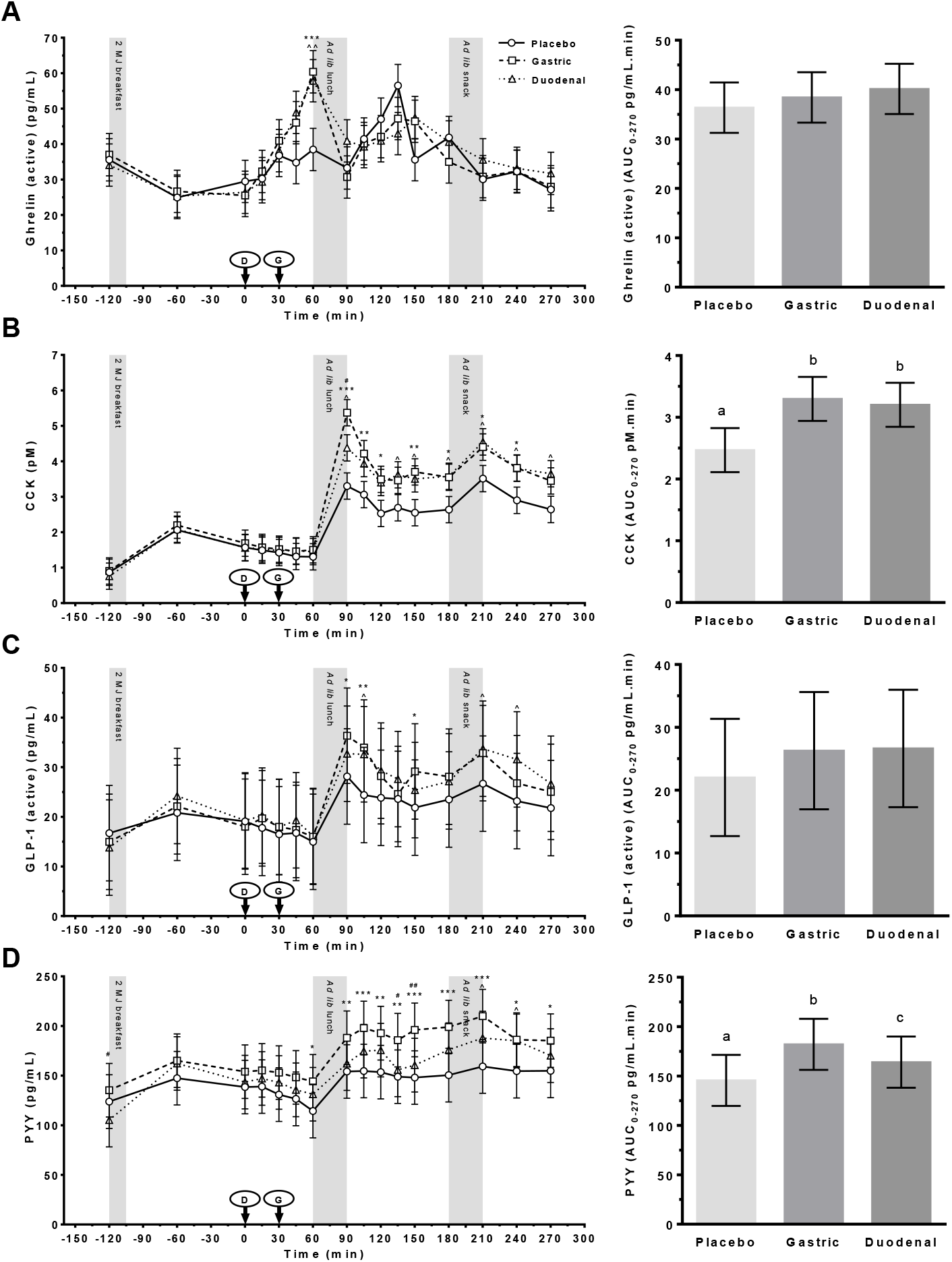
Plasma concentrations of (A) ghrelin (active); (B) cholecystokinin (CCK); (C) glucagon like peptide-1 (active) (GLP-1) and (D) peptide YY (PYY) following administration of a control (Placebo) or Amarasate^®^ targeted to either the small intestine (Duodenal) or stomach (Gastric) using delayed-release or standard capsules, respectively. Arrows indicate capsule administration; grey bars indicate the time allowed for the 2 MJ fixed energy breakfast and the *ad libitum* lunch and snack. Analysis was conducted using the mixed procedure (SAS 9.4) with treatment, time, visit number and treatment order as factors. Significant effects of treatment (B and D, p < 0.005) and a treatment x time interaction (A and C, p < 0.04) were observed. Fisher’s LSD *post hoc* pairwise comparisons: gastric v placebo (*p < 0.05, ** p < 0.01, *** p < 0.001); duodenal v placebo (^p < 0.05, ^^ p < 0.01, ^^^ p < 0.001); gastric v duodenal (^#^p < 0.05, ^##^ p < 0.01, ^###^ p < 0.001). Histograms show effect of treatment on AUC_0-270 min_ for each hormone from 0 to 270 min. Analysis was conducted using the mixed procedure (SAS 9.4) with treatment, visit number and treatment order as factors. A significant effect of treatment was observed for B and D (p = 0.001) only, with letters denoting significantly (p < 0.05) different means. Values are means ± SEM; n = 18. *Ad lib, ad libitum*.

##### Ghrelin

Plasma concentrations of the orexigenic hormone ghrelin exhibited a significant treatment x time interaction (F_30,440_ = 1.74, *p* = 0.010). Subsequent p*ost hoc* analysis demonstrated a significant increase in ghrelin in both gastric (p = 0.0009) and duodenal (p = 0.004) treatments compared with the placebo, immediately prior to the *ad libitum* lunch (T = 60). No significant differences were detected between any of the treatments at any post-lunch time point or for the AUC_0-270 min_ response (**Figure 3A**).

##### CCK

A significant main effect of treatment (F_2,70_ = 5.8, *p* < 0.005) and treatment x time (F_30,481_ = 1.9, *p* = 0.004) interaction were observed for plasma concentrations of the anorexigenic hormone CCK, with a similar pattern of enhanced postprandial CCK secretion observed in both the gastric and duodenal treatments compared with the placebo (**Figure 3B**). *Post hoc* analysis demonstrated that plasma CCK concentrations were significantly increased (p < 0.05) in both Amarasate^®^ treatments compared with the placebo at T = 90, 150, 180, 210, and 240 min. Significant increases (p < 0.05) were also seen at T = 105 and 120 min in the gastric and at T = 135 and 270 min in the duodenal treatments compared with the placebo. A significant difference between the gastric and duodenal treatments was observed at the 90-min time point only (*p* = 0.024).

A significant effect of treatment was also seen for the CCK AUC_0-270 min_ responses (F_2,32_ = 8.66, p = 0.001), with increased hormone secretion observed in both the duodenal (p = 0.002) and gastric (p < 0.001) treatments compared with the placebo (**Figure 3B**). Gastric and duodenal treatments did not differ significantly from each other.

##### GLP-1

Plasma concentrations of the insulin secretagogue and anorexigenic hormone GLP-1 exhibited considerable inter-individual variability (including one individual who exhibited approximately forty times average baseline levels), though a significant treatment x time interaction (F_30,422_ = 1.5, *p* = 0.038) was observed. *Post hoc* analysis demonstrated an enhanced (p < 0.05) postprandial response to the *ad libitum* lunch in the gastric treatment at T = 90, 105 and 150 min compared with the placebo (**Figure 3C**). The duodenal treatment elicited a similarly enhanced postprandial response, with significant (p < 0.05) increases at T = 105, 210 and 240 min compared with the placebo. Gastric and duodenal treatments did not differ significantly from each other at any time point. No evidence for a main effect of treatment was seen for GLP-1 AUC_0-270 min_ responses (**Figure 3C**).

##### PYY

A significant effect of treatment (F_2,35_ = 7.6, *p* = 0.002) and a treatment x time (F_30,475_ = 1.5, *p* = 0.042) interaction were observed for plasma concentrations of the anorexigenic gut hormone PYY (**Figure 3D**). Though considerable inter-individual variability was observed in baseline concentrations (two participants were ∼10 fold higher than average). *Post hoc* analysis demonstrated that when compared with the placebo, gastric delivery of Amarasate^®^ produced significant increases in PYY immediately prior to the lunch (T= 60), with differences becoming more apparent post lunch through to the end of the session (T = 90–270 min, *p* < 0.05). The PYY response to the duodenal treatment was significantly elevated relative to the response in the placebo treatment only after the *ad libitum* snack at T= 210 and 240 (p < 0.05) minutes. A significant difference between gastric and duodenal treatments was also observed at T = -120, 135 and 150 min (p < 0.05).

A significant effect of treatment was seen for the PYY AUC_0-270 min_ responses (F_2,32_ = 11.14, p < 0.001), with significantly increased PYY secretion observed in the gastric compared with both the duodenal (p = 0.027) and placebo (p <0.0001) treatments. PYY release was also significantly (p = 0.023) greater in the duodenal treatment than in the placebo (**Figure 3D**)

#### Glucose, Insulin, GIP and PP

Effects of treatment on plasma concentrations and AUC_0-270 min_ responses of glucose, insulin, GIP and PP are shown are shown in **Figure 4A–D**.

**Figure 4.**
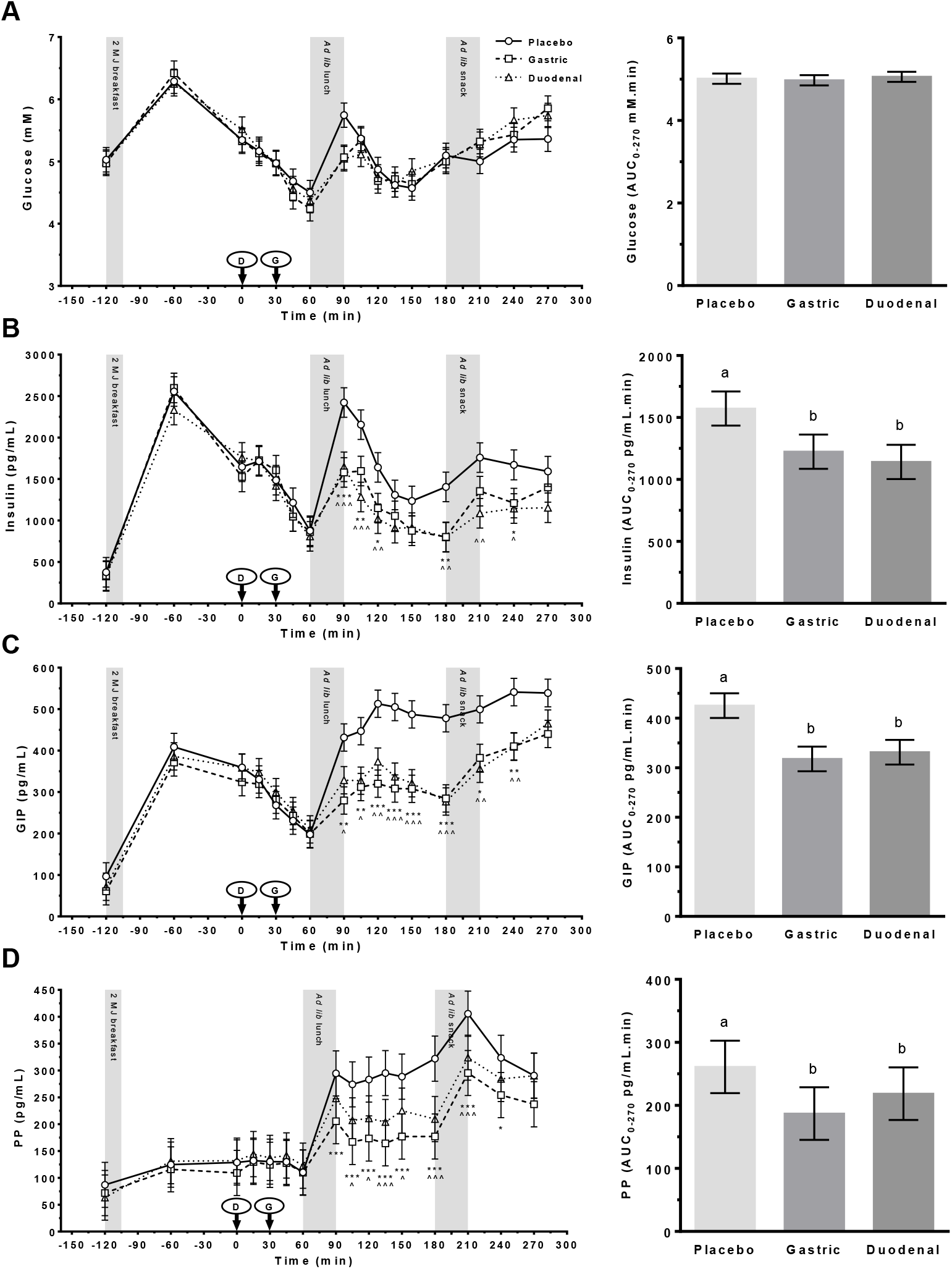
Plasma concentrations of (A) glucose; (B) insulin; (C) glucose-dependent insulinotropic polypeptide (GIP); and (D) pancreatic polypeptide (PP) following administration of a control (Placebo) or Amarasate^®^ targeted to either the small intestine (Duodenal) or stomach (Gastric) using delayed-release or standard capsules, respectively. Arrows indicate capsule administration; grey bars indicate the time allowed for the 2 MJ fixed energy breakfast and the *ad libitum* lunch and snack. Analysis was conducted using the mixed procedure (SAS 9.4) with treatment, time, visit number and treatment order as factors. A significant effect of treatment (B–D, p < 0.003) was observed. Fisher’s LSD *post hoc* pairwise comparisons: gastric v placebo (*p < 0.05, ** p < 0.01, *** p < 0.001); duodenal v placebo (^p < 0.05, ^^ p < 0.01, ^^^ p < 0.001); gastric v duodenal (^#^p < 0.05, ^##^ p < 0.01, ^###^ p < 0.001). Histograms show effect of treatment on AUC_0-270 min_ for each hormone from 0 to 270 min. Analysis was conducted using the mixed procedure (SAS 9.4) with treatment, visit number and treatment order as factors. A significant effect of treatment was observed for B–D (p < 0.001) only, with letters denoting significantly (p < 0.05) different means. Values are means ± SEM; n = 18. *Ad lib, ad libitum*.

##### Glucose

Changes in glucose with time (**Figure 4A**) suggest that Amarasate^®^ treatment modified postprandial hyperglycemia following the *ad libitum* lunch (T = 90) only. However, no significant main effect of treatment or treatment x time interaction was observed for blood glucose concentrations or in the glucose AUC_0-270 min_ response.

##### Insulin

Plasma insulin concentrations exhibited a significant effect of treatment (F_2,39_ = 8.6, *p* < 0.001) and a treatment x time (F_30,332_ = 1.8, *p* = 0.011) interaction (**Figure 4B**). *Post hoc* analysis demonstrated that insulin responses to the *ad libitum* lunch and snack showed a significant reduction (p < 0.05) following the gastric and duodenal treatments at T= 90, 105, 120, 180 and 210 min compared with the placebo. This difference extended out to T = 240 min for the gastric treatement only. Insulin responses in the gastric and duodenal treatments did not differ significantly from each other at any time point. A highly significant effect of treatment (F_2,32_ = 10.8, *p* = 0.0003) was also observed in insulin AUC_0-270 min_ responses (**Figure 4B**), with a reduction in postprandial insulin secretion following the gastric (p = 0.0013) and duodenal (p = 0.0001) treatments compared with the placebo. Insulin AUC_0-270 min_ responses in the gastric and duodenal treatments did not differ significantly from each other.

##### GIP

Plasma concentrations of the insulin secretagogue GIP exhibited a significant effect of treatment (F_2,65_ = 6.8, *p* < 0.002) and a treatment x time (F_30,517_ = 1.7, *p* = 0.010) interaction. *Post hoc* analysis demonstrated that the postprandial response to the *ad libitum* lunch and snack were significantly reduced in both the gastric and duodenal treatments compared with the placebo from T = 90 to 240 min (*p* < 0.050). Gastric and duodenal treatments did not differ significantly from each other at any timepoint. GIP AUC_0-270 min_ responses (**Figure 4C**) exhibited a highly significant effect of treatment (F_2,32_ = 15.5, p <0.0001), with reductions in postprandial GIP secretion following the gastric (p < 0.0001) and duodenal (p < 0.0001) treatments compared with the placebo. Gastric and duodenal treatments did not differ significantly from each other.

##### PP

Plasma concentrations of the pancreatic hormone PP exhibited a significant effect of treatment (F_2,69_ = 8.8, *p* = 0.0004) and a treatment x time interaction (F_30,491_ = 1.91, p = 0.003), increasing following meals in all treatment groups (**Figure 4D**). *Post hoc* analysis demonstrated that postprandial PP responses were significantly (p < 0.05) reduced in both the gastric (T = 90–240 min) and duodenal (T = 105–210 min) treatments compared with the placebo. Duodenal and gastric treatments did not differ significantly from each other at any timepoint.

A highly significant effect of treatment (F_2,32_ = 11.6, p = 0.0002) was seen for the PP AUC_0-270 min_ responses, with reduced hormone secretion observed in both the duodenal (p = 0.0096) and gastric (p < 0.0001) treatments compared with the placebo (**Figure 4D**). Gastric and duodenal treatments did not differ significantly from each other.

#### VAS – appetite

Effects of treatment on the subjective ratings of hunger, fullness, prospective consumption, satiety and thirst over time and as AUC_0-300 min_ are shown in **Figure 5A–E**. A predictable pattern driven by meal timing was seen in all VAS profiles. However, there was no evidence for a significant main effect of treatment or treatment x time interaction for any of the changes in VAS appetite profiles or in AUC_0-300 min_.

**Figure 5.**
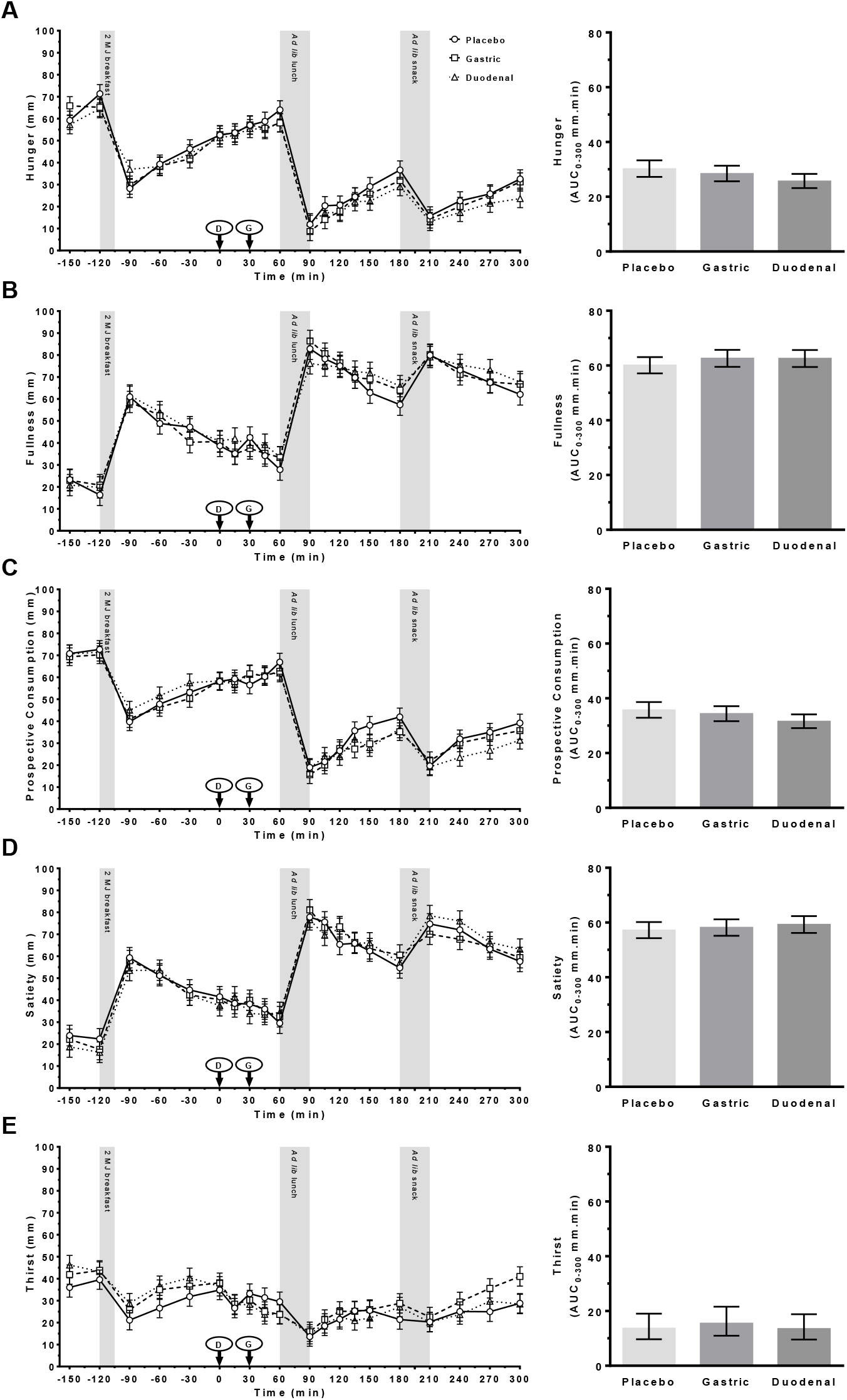
VAS ratings of (A) hunger; (B) fullness; (C) prospective consumption; (D) satiety; and (E) thirst following administration of a control (Placebo) or Amarasate^®^ targeted to either the small intestine (Duodenal) or stomach (Gastric) using delayed-release or standard capsules, respectively. Arrows indicate capsule administration; grey bars indicate the time allowed for the 2 MJ fixed energy breakfast and the *ad libitum* lunch and snack. Analysis was conducted using the mixed procedure (SAS 9.4) with treatment, time, visit number and treatment order as factors. No main effect of treatment or a treatment x time interaction was observed for any measure. Histograms show mean AUC_0-300 min_ for each VAS measure from 0 to 300 min. Analysis was conducted using the mixed procedure (SAS 9.4) with treatment, visit number and treatment order as factors. No significant effects of treatment were seen. Values are means ± SEM; n = 19. *Ad lib, ad libitum*.

#### VAS – vitality

Effects of treatment on subjective ratings of energy and relaxation are shown in **Supplemental Figure 3A–B**. Ratings of energy exhibited a significant treatment effect (F_2,98_= 3.29, *p* = 0.041), with *post hoc* analysis demonstrating significantly (p < 0.050) lower energy ratings in the duodenal treatment at T = 120, 135, 150 and 270 min, and in the gastric treatment at T = 150 and 270 min, compared with the placebo. A signicant treatment effect (F_2,32_ = 4.65, p = 0.017) was also seen for AUC_0-300 min_ responses, with lower ratings for the gastric (p = 0.019) and duodenal (p = 0.009) treatments compared with the placebo (**Supplemental Figure 3A**). Subjective ratings of relaxation were similar throughout the day, with no significant main effects of treatment or treatment x time interactions (**Supplemental Figure 3B**).

#### VAS – Gastrointestinal discomfort

Effects of treatment on subjective ratings of nausea, urge to vomit, bloating, abdominal discomfort and heartburn are shown in **Figure 6A–E**.

**Figure 6.**
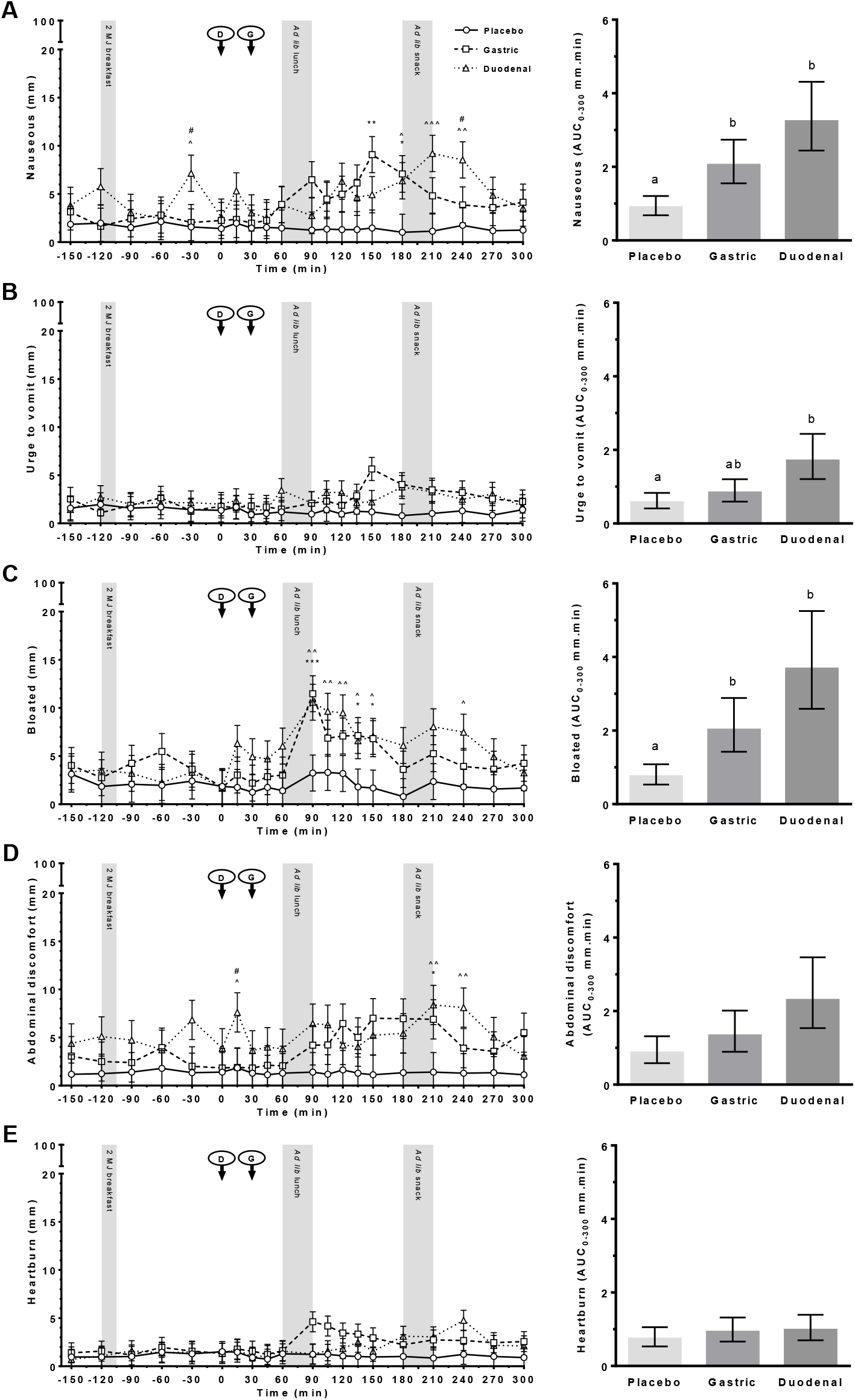
VAS ratings of (A) nausea; (B) urge to vomit; (C) bloating; (D) abdominal discomfort; and (E) heartburn following administration of a control (Placebo) or Amarasate^®^ targeted to either the small intestine (Duodenal) or stomach (Gastric) using delayed-release or standard capsules, respectively. Arrows indicate capsule administration; grey bars indicate the time allowed for the 2 MJ fixed energy breakfast and the *ad libitum* lunch and snack. Analysis was conducted using the mixed procedure (SAS 9.4) with treatment, time, visit number and treatment order as factors. A significant effect of treatment was observed for A, C and D (p < 0.010). Fisher’s LSD *post hoc* pairwise comparisons: gastric v placebo (*p < 0.05, ** p < 0.01, *** p < 0.001); duodenal v placebo (^p < 0.05, ^^ p < 0.01, ^^^ p < 0.001); gastric v duodenal (^#^p < 0.05). Histograms show effect of treatment on AUC_0-300 min_ for each VAS scale from 0 to 300 min. Analysis was conducted using the mixed procedure (SAS 9.4) with treatment, visit number and treatment order as factors. A significant effect of treatment was observed for A–C (p < 0.05), with letters denoting significantly (p < 0.05) different means. Values are means ± SEM; n = 19. *Ad lib, ad libitum*.

##### Nausea

Ratings of nausea (**Figure 6A**) exhibited a significant treatment effect (F_2,124_ = 5.5, p = 0.005), with *post hoc* analysis demonstrating significantly (p < 0.050) higher nausea ratings in the duodenal treatment at T = -30, 180, 210 and 240 min, and in the gastric treatment at T = 150 and 180 min, compared with the placebo. Signifcant differences between the gastric and duodenal treatments were seen at T = -30 and 240 min. A significant treatment effect (F_2,32_ = 10.9, p = 0.0002) on AUC_0-300 min_ responses was also observed, with lower ratings in the gastric (p = 0.006) and duodenal (p < 0.0001) treatments than in the placebo.

##### Urge to vomit

There was no evidence for a significant main effect treatment or treatment x time interactions in the urge to vomit (**Figure 6B**). However, a significant treatment effect (F_2,32_ = 3.95, p = 0.029) on AUC_0-300 min_ responses was seen, with a small increase in the urge to vomit (p = 0.009) in the duodenal treatment compared with the placebo.

##### Bloating

A significant main effect of treatment (F_2,127_ = 6.83, p = 0.002) was seen for subjective ratings of abdominal bloating (**Figure 6C**). *Post hoc* analysis demonstrated significantly (p < 0.050) higher ratings of abdominal bloating in the duodenal treatment at T = 90–150 and 240 min, and in the gastric treatment at T = 90, 135 and 150 min, than in the placebo (**Figure 6C**). A significant treatment effect (F_2,32_ = 7.8, p = 0.002) was also seen on AUC_0-300 min_ responses, with a small increase in bloating in the duodenal (p = 0.001) and gastric (p = 0.020) treatments compared with the placebo.

##### Abdominal discomfort

A significant main effect of treatment (F_2,116_ = 5.65, p = 0.005) was seen for ratings of abdominal discomfort (**Figure 6D**), with *post hoc* analysis demonstrating significantly (p < 0.050) higher abdominal discomfort ratings in the duodenal treatment at T = 15, 210 and 240 min, and in the gastric treatment at T = 210 min, than in the placebo. Signifcant differences between gastric and duodenal treatments were seen at T = 15 min. However, there was no evidence for a significant effect of treatment on AUC_0-300 min_.

##### Heartburn

No significant main effects of treatment or treatment x time interactions were observed for ratings of heartburn or AUC_0-300 min_ response (**Figure 6E**).

#### VAS – Meal palatability

There was no evidence for a main effect of treatment on VAS ratings of pleasantness, visual appeal, smell, taste, aftertaste, or overall palatability for the fixed-energy breakfast or the *ad libitum* lunch and snack outcome meals (**Supplemental Figure 4**).

#### Profile of mood states (POMS)

The effects of treatment on the six mood subscales and total mood disturbance measured prior to treatment administration (Pre) and in the POMS questionnaire are shown in **Supplemental Figure 5A–G**. Only depression-dejection (F_2,83_ = 3.4, p = 0.038) and anger-hostility (F_2,83_ = 6.14, p = 0.003) exhibited a significant main effect of treatment. *Post hoc* analysis demonstrated a small but significant (p = 0.011) increase (2.3 ± 1.2) in scoring of depression-dejection following the gastric treatment compared with the placebo (1.5 ± 1.2) (**Supplemental Figure 5B**). Small but significant increases in ratings of anger-hostility were also seen following the gastric (2.2 ± 1.2) treatment compared with the duodenal (1.5 ± 1.2, p = 0.016) and placebo (1.4 ± 1.2, p = 0.004) treatments (**Supplemental Figure 5C**). However, these small changes in mood state may be a reflection of social and envrionmental interactions e.g. self-entertainment activities such as watching TV, reading and socialising.

#### Adverse symptoms

The numbers of participants reporting adverse symptoms such as loose stool/diarrhea, nausea, rumbling or upset stomach, bloating and headache during the study day, and their subjective ratings of severity (mild, moderate or severe) are shown in **Table 2**. The primary analysis of all 19 participants revealed a total of 14 adverse symptoms, the majority of which (93%) occurred while on the gastric treatment. No adverse symptoms were reported while on the placebo treatment and only one individual reported a reduced frequency of defecation in the week following the duodenal treatment (washout period), which may not have been attributable to the treatment, given the delay. The participant excluded for failure to comply with the study protocol experienced moderate-intensity loose stools and nausea in both gastric and duodenal treatments and was included in a separate analysis (in brackets) of 20 participants for completeness.

**Table 2.** The effects of treatment on numbers of reported adverse symptoms and range of self-reported intensities^1^.

|  | Placebo | Gastric | Duodenal |
| --- | --- | --- | --- |
| **Nausea** | - | 2/19 (3/20)<sup>2</sup> mod | -(1/20)<sup>2</sup>mod |
| **Loose stool/diarrhea** | - | 6/19 (7/20)<sup>2</sup> mild-mod | -(1/20)<sup>2</sup>mod |
| **Stomach rumbling** | - | 1/19 mild | - |
| **Upset stomach** | - | 1/19 mild | - |
| **Bloating** | - | 2/19 mod | - |
| **Headache** | - | 1/19 mild | - |
| **Frequency of defecation** | - | - | 1/19 mod<sup>3</sup> |

<sup>1</sup> Severity of adverse events was reported using a three-point scale of mild, moderate (mod) or severe over the study visit and washout period for each treatment. Treatments comprised a vehicle control (Placebo) or Amarasate<sup>®</sup> targeted to either the small intestine (Duodenal) or stomach (Gastric) using delayed-release or standard capsules, respectively. <sup>2</sup>The participant excluded for failure to comply with the study protocol experienced moderate-intensity loose stools and nausea in both gastric and duodenal treatments and was included in the adverse symptom reporting as a separate analysis (in brackets) with all n=20 participants. <sup>3</sup>One participant noted a reduced frequency of defecation over the following week washout period.

## 4. Discussion

Gastrointestinal delivery of a bitter hop extract significantly decreased energy intake and increased appetite-suppressing CCK, PYY and GLP-1 plasma concentrations. These changes occurred without significant effects on subjective measures of appetite or the hedonic properties of the test meals. However they were accompanied by small increases in subjective ratings of nausea, bloating, urge to vomit and abdominal discomfort, all of which are expected to decrease EI, and may be confounders. The magnitude of total EI suppression (18%) is significant in the context of weight management applications (67) and compares favourably with results from previous studies in humans (0–22%) that have used either encapsulation, intragastric or intraduodenal delivery of a variety of bitter tastants (38, 40, 42, 43, 46, 68, 69).

The current study supports a mechanism of action involving enhanced and sustained release of the anorexigenic gut hormones CCK, GLP-1 and PYY from intestinal EECs. All three gut peptide hormones play a key role in the homeostatic regulation of energy intake, appetite and GI function (reviewed in (70)) and have previously been shown to respond to T2R ligands (29, 30, 44, 48). Maximum post-prandial increases in CCK following Amarasate^®^ treatments were almost 6-fold that of baseline and in the upper range reported for dietary interventions (0.5–7.9 fold) (71)). Greater variablity between individuals and smaller fold changes were observed for GLP-1 (2.4 fold) and PYY (1.8 fold). A recent meta-analysis of CCK, GLP-1 and PYY infusion studies (71) that proposed that the minimum fold changes required to decrease *ad libitum* energy intake were 3.6, 4.0 and 3.1-fold, respectively.

A significant enhancement of the orexigenic hormone ghrelin response prior to the lunch was also seen for both gastric and duodenal targetting of the hop extract. This is consistent with the duodenum being a source of ghrelin secretion, second only to the stomach (72, 73), although pyloric reflux may also play a role (74). Gavage of T2R agonists has also been shown to stimulate the secretion of ghrelin in mice, resulting in a temporary increase in food intake (75). However, our results contrast with several recent reports of either unchanged or supressed ghrelin following intragastric infusion of T2R agonists (quinine, denatoium benzoate) in humans (41, 76), indicating potential T2R specificity in this response. The mechanism(s) by which T2R agonists stimulate ghrelin secretion are poorly understood, as gastric ghrelin-secreting cells are of the closed type and do not directly contact the GI lumen.

It is also noteworthy that there was no significant treatment-induced difference in VAS measurements relating to appetite despite the significant decrease in energy intake seen with both hop treatments. Although correlations between subjective assessments (e.g. hunger) and behavioural effects (e.g. energy intake) are often observed, they assess fundamentally different things, have been reported to show weak correlations, and do not always concur (77, 78). Previous studies using either gastric or duodenal delivery of T2R agonists have shown effects on subjective measures of appetite in both men (79-81), and women (41, 82), although many studies show no response (43-46). Interestingly, particpants in the current study did achieve similar feelings of fullness at the *ad libitum* test meals after consuming less food when taking both hop treatments compared with the placebo. Viewed in this context, Amarasate^®^ treatment may modulate early satiety, which is associated with impaired gastric accommadation and gastric emptying (83).

Glucoregulatory hormones (e.g. GLP-1, GIP, insulin) and the slowing of gastric emptying are key determinants of postprandial glycemia. Bitter tastants have been shown to stimulate the secretion of the incretin hormone GLP-1 from EEC cell lines (30, 48), while in mice gavage of bitter gourd extract (84) or denatonium benozate (30) stimulates GLP-1 and subsequent insulin secretion, leading to lowering of blood glucose. A recent study in healthy men also demonstrated that intragastric and intraduodenal administration of the bitter tastant quinine similarly lowered plasma glucose, increased plasma insulin and GLP-1, and slowed gastric emptying (40). The current data also demonstrate an enhancement in the postprandial GLP-1 response following gastric and duodenal targeting of hop extract. However, this response was accompanied by a substantial reduction in the postprandial insulin response, with little change in plasma glucose compared with that in the placebo. Interestingly, GIP, the only gut peptide hormone measured that is secreted from the enteroendocrine K-cell subtype, also exhibited a substantial reduction in postprandial response following hop treatments. This is contrast to the observed stimulation of CCK, GLP-1 and PYY producing EECs, suggesting that K-cells lack the appropriate T2Rs.

GIP has been shown to be responsible for the majority of the incretin effect in healthy subjects, affecting glycemic levels during the whole postprandial period (85). In contrast, GLP-1 primarily affects glycaemic regulation in the early postprandial phase, delaying gastric emptying and reducing plasma glucagon levels (85). GIP has also recently been demonstrated as a PP secretagogue (85, 86). Hence the suppression of postparndial GIP in the hop treatment groups may in part explain the suppression of insulin and PP observed.

GIP secretion is driven primarliy by the rate of macronutrient delivery from the stomach to the duodenum (i.e. rate of gastric emptying) (87). Any delay in gastric emptying in the absence of a treatment induced-stimulation would potentially result in this observed decrease (88-90). Although this study did not measure gastric emptying *per se*, an established action of CCK, GLP-1 and PYY is to delay gastric emptying (91-93). Importantly, regulation of postprandial glycemia was maintained despite reductions in GIP and insulin, indicating a metabolic shift towards greater insulin sensitivity, a possible consequence of increased GLP-1 secretion. Replication of these results using a fixed energy meal would be ideal, as this would remove any influence from the inter-treatment differences in absolute energy intake that occurred at the *ad libitum* meals.

Off-target effects of Amarasate^®^ treatments included small (<10 mm) but significant increases in subjective ratings of nausea, bloating and abdominal discomfort, which are consistent with known effects of CCK, GLP-1 and PYY on upper gastrointestinal sensations (94, 95). The known sedative activity of hop bitter acids may also have contributed to the small decline in subjective ratings of energy following Amarasate^®^ treatment (96, 97), although no corresponding effect on relaxation was observed. Virtually all reported adverse symptoms were associated with gastric targeting of the hops extract, with 32% of participants in this treatment reporting an acute bout of mild- to moderate-intensity diarrhea. Targeting delivery to the small intestine improved tolerance of the hop treatment, suggesting that gastric

T2Rs may play a key role in detection of ingested toxins, stimulating a host defence mechanism involving net secretion of fluid and electrolytes into the intestinal lumen, accelerating intestinal transit to flush harmful compounds from the GI tract in a process similar to that described for T2Rs in the human and rat large intestine (98). Further optimisation of the dosage of hops extract used and its timing relative to meals may also contribute to a reduction in the side-effect profile.

The Amarasate^®^ extract used in the current study is a supercritical CO_2_ extract of hop containing a number of hop bitter acids (e.g. cohumulone, humulone, adhumulone, colupulone, lupulone and adlupulone). These α- and β-acids are potent ligands for hT2R-1, 14 and 40, exhibiting reported thresholds of activation as low as 3 nM (47). All three hop-responsive hT2Rs have previously been identified in either the small (31) or large intestine (99, 100). However, little is known regarding the profile of hT2Rs expression in specific EEC cell-types. The functional data from the current study would suggest CCK, GLP-1, PYY and ghrelin-producing EECs express T2R-1, 14 or 40, a T2R expression profile not shared by GIP-producing EECs.

It is worth noting that other compounds derived from hops acids have previously been examined as anti-obesity targets, and potential exists for overlapping or synergistic mechanisms of action. Oxidised hop bitter acid extracts have been reported by Morimoto-Kobayashi et al. to reduce fat mass in an overweight population through increased brown adipose tissue thermogenesis (55, 56), with recent work in rats suggesting CCK secretion as a potential mediator of this bioactivity (32). Isomerised α-acids and their derivatives have also shown anti-obesogenic effects, putatively *via* metabolic regulation (39, 50), although a recent report in mice highlighted a possible role for the T2Rs and anorexigenic hormones (48). Kok et al. (48) showed that the synthetic substituted 1,3-cyclopentadione isomerised alpha acid derivative KDT501 (KinDex Pharmaceuticals) activated T2R1 *in vitro* and when administered to mice resulted in increased GLP-1, CCK and ghrelin plasma concentrations, and improved glucose and insulin responses.

Some limitations of our study should be noted. Targeted delivery of Amarasate^®^ to the duodenum may not have occurred in all cases, as the press-fit delayed-release capsules used, can leak (101) or disassemble (102) under gastric conditions *in vitro*. In addition, the short intervals between the *ad libitum* lunch, snack and end of daily monitoring may have prevented appropriate treatment differences developing in appetitive VAS measures such as hunger, fullness and prospective consumption (57). The study could also have benefited from the inclusion of measures of gastric emptying. However, GIP secretion is dependent upon nutrient delivery to the duodenum (103) and indirectly supports delayed gastric emptying as a mechanism of action. Another limitation of the study was our inclusion of only healthy-weight males as paticipants, done to exclude the potentially confounding effects of the menstrual cycle on energy intake (104) and to ensure robust appetite and glycaemic regulatory mechanisms (9-13). Finally, the effect of repetitive or chronic administration of hop extract on appetite regulation, including possible compensatory mechanisms and effects on weight management, are unknown. Thus, further long-term studies are warranted.

In conclusion, both gastric and duodenal delivery of Amarasate^®^, a bitter hop extract, suppressed total EI and enhanced pre-meal ghrelin and postprandial CCK, GLP-1 and PYY responses, providing a potential “bitter brake” on energy intake in healthy-weight males. These changes occurred without significant effects on subjective measures of appetite or on any measure of the hedonistic properties of the test meals though small but significant increases were observed in some measures of gastrointestoinal discomfort. Changes in glycaemic regulation were also observed, with reductions in postprandial insulin, PP and GIP responses without a significant effect on the glycaemic response to the *ad libitium* test meals.

These data highlight the potential of hop compounds as novel therapeutics to regulate both acute energy intake and glycaemic regulation via modulation of gut peptide release and delayed gastric emptying. Further studies investigating the longer-term effects of lower doses of hop extract as a tool for improved weight management and glycemic regulation should be conducted, to determine potential efficacy.

## Data Availability

Data described in the manuscript, code book, and analytic code will be made available upon request

## 6. Acknowledgements

The authors thank the individuals who kindly agreed to be participants in the clinical trial. We would also like to thank the nurse practitioners who assisted with blood sampling during the clinical trial, Dr Raina Wong for assistance with hormone analysis, Dr Ron Beatson and Dave Anderson for their assistance with hops analysis and selection, and Dr Russell Walmsley for providing medical oversight for this trial. We also thank Prof. Richard Newcomb, Dr Roger Harker and Dr Pramod Gopal for review of the manuscript. This research was funded by the New Zealand Government (NZ Ministry of Business, Innovation & Employment, contract C11X1004).

## 7. Author Contributions

Contributor roles (Credit, CASRAI): conceptualization and supervision: JI; funding acquisition: KS, JI; methodology: EW, JI and SP; project administration EW; investigation: EW, KL, MP, HS, CL, KS and JI; data curation: JI; formal analysis MW; visualisation: EW and JI ; Writing – original draft: EW, SP and JI; Writing – review & editing: All Authors.

## Abbreviations list

(AUC): Area under the curve
(T2R): bitter taste receptor
(CO_2_): carbon dioxide
(CCK): cholecystokinin
(EI): energy intake
(EEC): enteroendocrine cell
(GI): gastrointestinal
(GLP-1): glucagon like peptide-1
(GIP): glucose-dependent insulinotropic polypeptide
(HbA1c): glycated hemoglobin
(hT2R): human bitter taste receptor
(LSD): least significant difference
(PP): pancreatic polypeptide
(PYY): peptide YY
(POMS): profile of mood states
(VAS): visual analogue scale

