## Supplemental tables and figures for "An extract of hops (*Humulus lupulus* L.) modulates gut peptide hormone secretion and reduces energy intake in healthy weight men: a randomised, cross-over clinical trial"

### Supplemental data

**Supplemental Table 1**. Treatment capsule allocation and timing of administration.

| Treatment | Delayed release capsule[1] (1100h, T = 0) | Standard capsule[2] (1130h, T = 30) |
|---|---|---|
| **Gastric** | 2x Vehicle control (125 mg oil[3]/capsule) | 2x Amarasate (250 mg extract[4]/capsule) |
| **Duodenum** | 2x Amarasate (250 mg extract[4]/capsule) | 2x Vehicle control (125 mg oil[3]/capsule) |
| **Placebo** | 2x Vehicle control (125 mg oil[3]/capsule) | 2x Vehicle control (125 mg oil[3]/capsule) |

[1]Delayed-release HPMC capsules (size 0, DRCaps™, Capsugel); [2]Standard HPMC capsules (size 0, VCaps™, Capsugel); [3]Canola oil; [4]Amarasate®, a bitter supercritical $CO_2$ extract of hops (*Humulus lupulus*).

**Supplemental Table 2.** Nutritional composition of the standard breakfast and *ad libitum* lunch and snack meals[1].

| | Weight (g) | Energy (kJ) | CHO (g) | CHO en% | Fat (g) | Fat en% | Protein (g) | Protein en% |
|---|---|---|---|---|---|---|---|---|
| **Standard breakfast (250 g serving, 2 MJ)** | | | | | | | | |
| Puffed rice cereal | 50 | 815 | 43 | 88 | 0.6 | 3 | 3.7 | 8 |
| Milk, low fat | 120 | 232 | 5.8 | 40 | 1.7 | 27 | 4.2 | 31 |
| White bread | 55 | 550 | 23.7 | 72 | 1.3 | 9 | 5 | 15 |
| Margarine, spread | 10 | 240 | 0 | 0 | 6.5 | 100 | 0 | 0 |
| Berry jam | 15 | 170 | 9.7 | 95 | 0 | 1 | 0.1 | 1 |
| ***Ad lib* lunch: pasta & meat sauce (2145 g serving, 8.9 MJ)** | | | | | | | | |
| Pasta, spirals, boiled | 981 | 5668 | 270 | 81 | 5.4 | 4 | 44 | 13 |
| Meat sauce, beef and tomato | 1164 | 3242 | 46 | 24 | 33 | 37 | 69 | 36 |
| ***Ad lib* snack: ham sandwich (348 g serving, 3.2 MJ)** | | | | | | | | |
| White bread | 204 | 2051 | 88 | 72 | 5 | 9 | 19 | 15 |
| Margarine, spread | 24 | 626 | 0 | 0 | 17 | 100 | 0 | 0 |
| Ham, sliced | 120 | 528 | 2 | 7 | 6 | 43 | 16 | 50 |

[1]Ad lib, *ad libitum*; CHO, carbohydrate; Prot, protein; En%, energy percentage.

**Supplemental Table 3.** Visual analogue scale questions and their left and right anchor statements used to access subjective ratings of the factors appetite, vitality, gastrointestinal discomfort and meal palatability.

| Factors | Question | Left Anchor | Right Anchor |
|---|---|---|---|
| **Appetite** | | | |
| Hunger | How hungry do you feel? | I am not hungry at all | I am as hungry as I've ever been |
| Fullness | How full do you feel? | I am not full at all | I am totally full |
| Satiety | How satisfied do you feel? | I am completely empty | I cannot eat another bite |
| Prospective Consumption | How much do you think you can eat now? | Nothing at all | A large amount |
| Thirst | How thirsty do you feel? | I'm not thirsty at all | I am as thirsty as I've ever been |
| **Vitality** | | | |
| Energetic | How energetic do you feel? | I do not feel energetic at all | I am as energetic as I have ever been |
| Relaxation | How relaxed do you feel? | I do not relaxed at all | I am as relaxed as I have ever been |
| **Gastrointestinal Discomfort** | | | |
| Urge to vomit | Do you have the urge to vomit? | Nothing at all | A large amount |
| Nauseous | Do you feel sick or nauseous? | Not at all | Very nauseous |
| Bloating | Do you feel bloated? | Not at all | A large amount |
| Abdominal discomfort | Do you have abdominal cramp or discomfort? | Nothing at all | A large amount |
| Heartburn | Do you have any heartburn? | Nothing at all | A large amount |
| **Meal palatability** | | | |
| Pleasantness | How pleasant was the meal? | Not at all pleasant | As pleasant as I have ever tasted |
| Visual Appeal | How did the meal look? | Bad | Good |
| Smell | How did the meal smell? | Bad | Good |
| Taste | How did the meal taste? | Bad | Good |
| Aftertaste | How much aftertaste was there? | None | Much |
| Overall Palatability | How appealing was the meal? | Bad | Good |

**Supplemental Table 4.** Immunoassay quality control values for the Magpix multiplexed magnetic bead assays[1] and the Cholecystokinin (CCK) radioimmunoassay[2].

| | Ghrelin (active) (pg/mL) | GIP (pg/mL) | GLP-1 (active) (pg/mL) | Insulin (pg/mL) | PP (pg/mL) | PYY (pg/mL) | CCK (pM) |
|---|---|---|---|---|---|---|---|
| Standard curve | 13.7–10,000 | 1.4–1000 | 2.7–2000 | 137–100,000 | 2.7–2000 | 13.7–10000 | 0.78–50 |
| Min DC | 13.6 | 0.7 | 1.4 | 58.9 | 1.8 | 11.4 | 0.63 |
| Max DC | 11956 | 2398 | 4595 | 231314 | 4094 | 6846 | 181 |
| Interassay %CV | 5.3 | 4.7 | 8.1 | 3.9 | 4.4 | 4.9 | 9.2 |
| Intraassay %CV | 2.5 | 3.4 | 3.9 | 3.1 | 3.8 | 6.3 | 4.1 |

[1]Plasma active ghrelin, active glucagon-like peptide-1 (GLP-1), total peptide YY (PYY), insulin, total glucose-dependent insulinotropic polypeptide (GIP) and pancreatic polypeptide (PP) concentrations were measured using a multiplexed magnetic bead assay (HMHMAG-34K; Merck-Millipore, Massachusetts USA). Cholecystokinin was measured using a commercial radioimmunoassay kit (EURIA-CCK, Eurodiagnostica). Minimal detectable concentration (Min DC), maximal detectable concentration (Max DC).

A

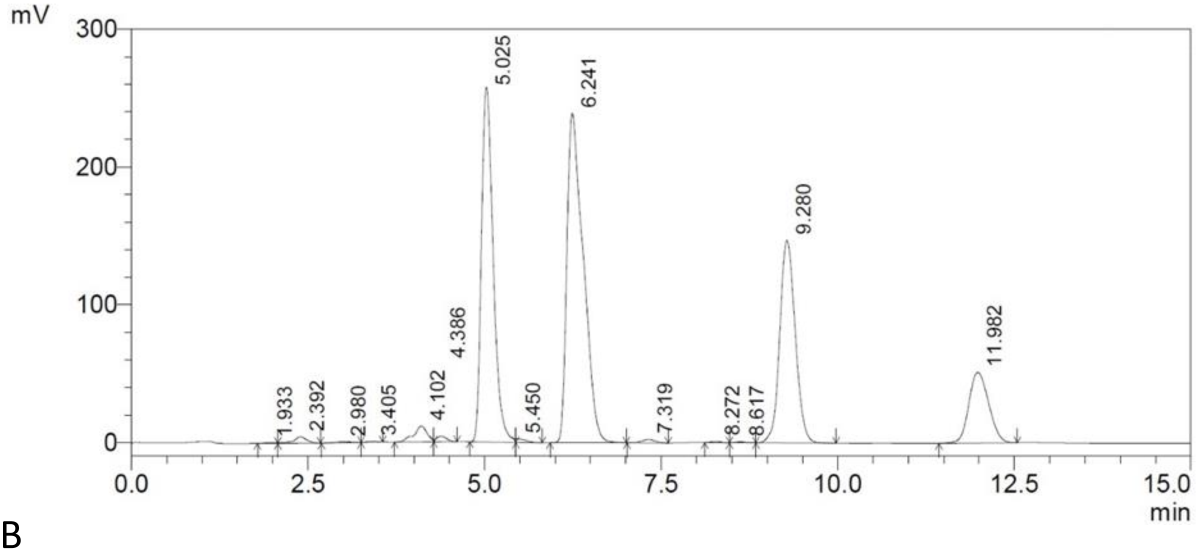


B

| Compound | Retention Time (RT) | Percentage |
|---|---|---|
| Xanthohumol | 2.392 | 0.07 |
| Cohumulone | 5.025 | 20.03 |
| Adhumulone/humulone | 6.241 | 30.96 |
| Colupulone | 9.280 | 20.25 |
| Adlupulone/lupulone | 11.982 | 9.20 |
| Total α-Acid | | 50.99 |
| Total β-Acid | | 29.45 |

**Supplemental Figure 1**. HPLC UV chromatogram (A) of the supercritical $CO_2$ hops extract used in the formulation of the Amarasate® treatments. Peak identifications, retention time and relative concentration expressed as percentages are listed in the included table (B).

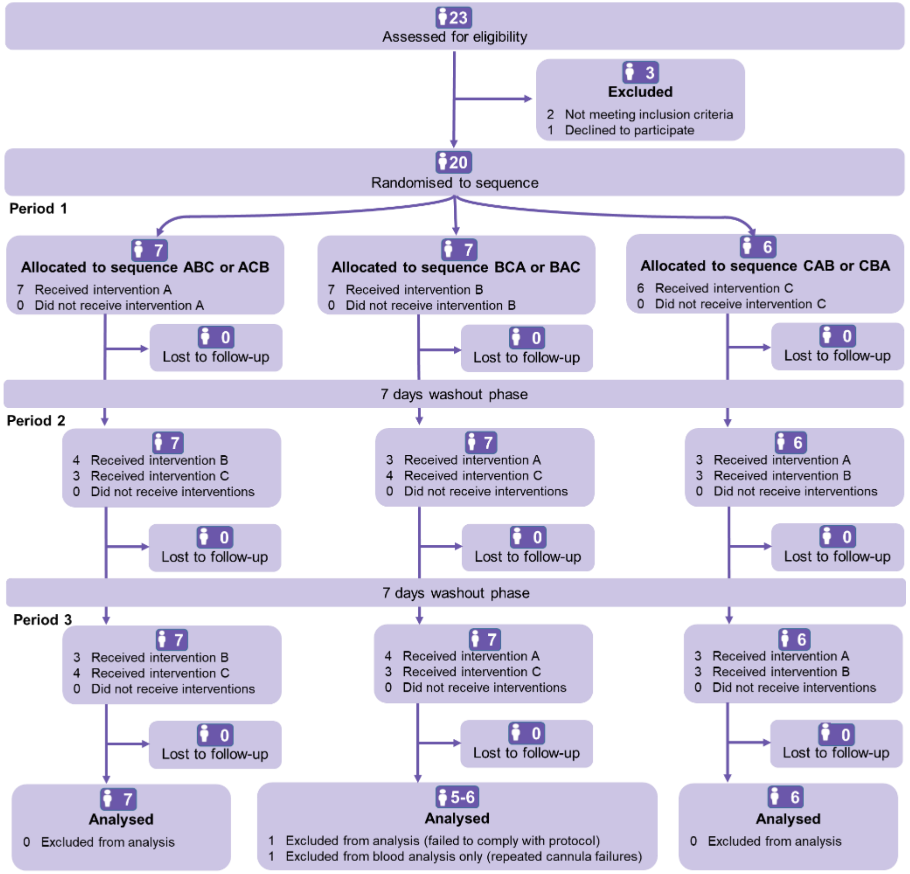


**Supplemental Figure 2.** CONSORT flow diagram showing the flow of participants through the process of recruitment, randomization to the six possible treatment sequences, exclusions and data analysis. Treatments are (A) duodenal and (B) gastric delivery of a hops extract or (C) placebo (vehicle control). Figure is based on the recommendations of the CONSORT extension to randomized crossover trials (105).

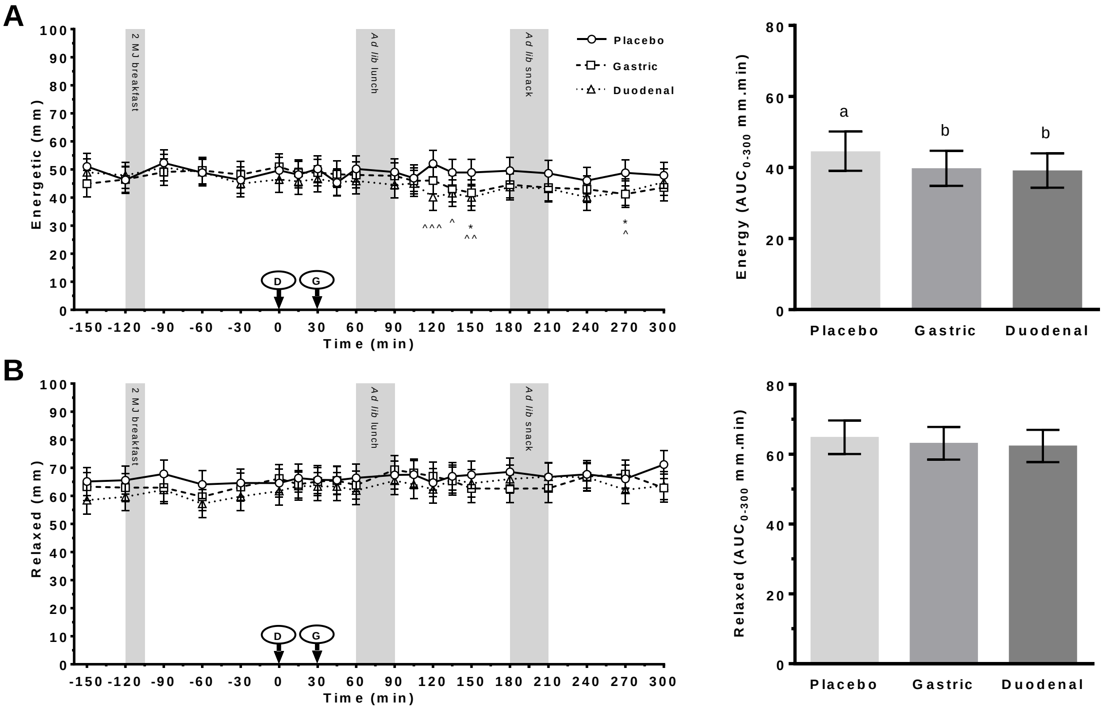


**Supplemental Figure 3** VAS ratings of (A) energetic and (B) relaxed following administration of a control (Placebo) or Amarasate® targeted to either the small intestine (Duodenal) or stomach (Gastric) using delayed-release or standard capsules, respectively. Arrows indicate capsule administration; grey bars indicate the time allowed for the 2 MJ fixed energy breakfast and the *ad libitum* lunch and snack. Analysis was conducted using the mixed procedure (SAS 9.4) with treatment, time, visit number and treatment order as factors. A significant effect of treatment ($p < 0.05$) was observed for energetic. Fisher's LSD *post hoc* pairwise comparisons: gastric v placebo (*$p < 0.05$); duodenal v placebo (^$p < 0.05$, ^^ $p < 0.01$, ^^^ $p < 0.001$). Histograms show effect of treatment on VAS AUC$_{0\text{-}300\text{ min}}$ for each vitality scale from 0 to 300 min. Analysis was conducted using the mixed procedure (SAS 9.4) with treatment, visit number and treatment order as factors. A significant effect of treatment was observed for energetic ($p = 0.017$), with letters denoting significantly ($p < 0.05$) different means. Values are means ± SEM; n = 19. *Ad lib*, *ad libitum*

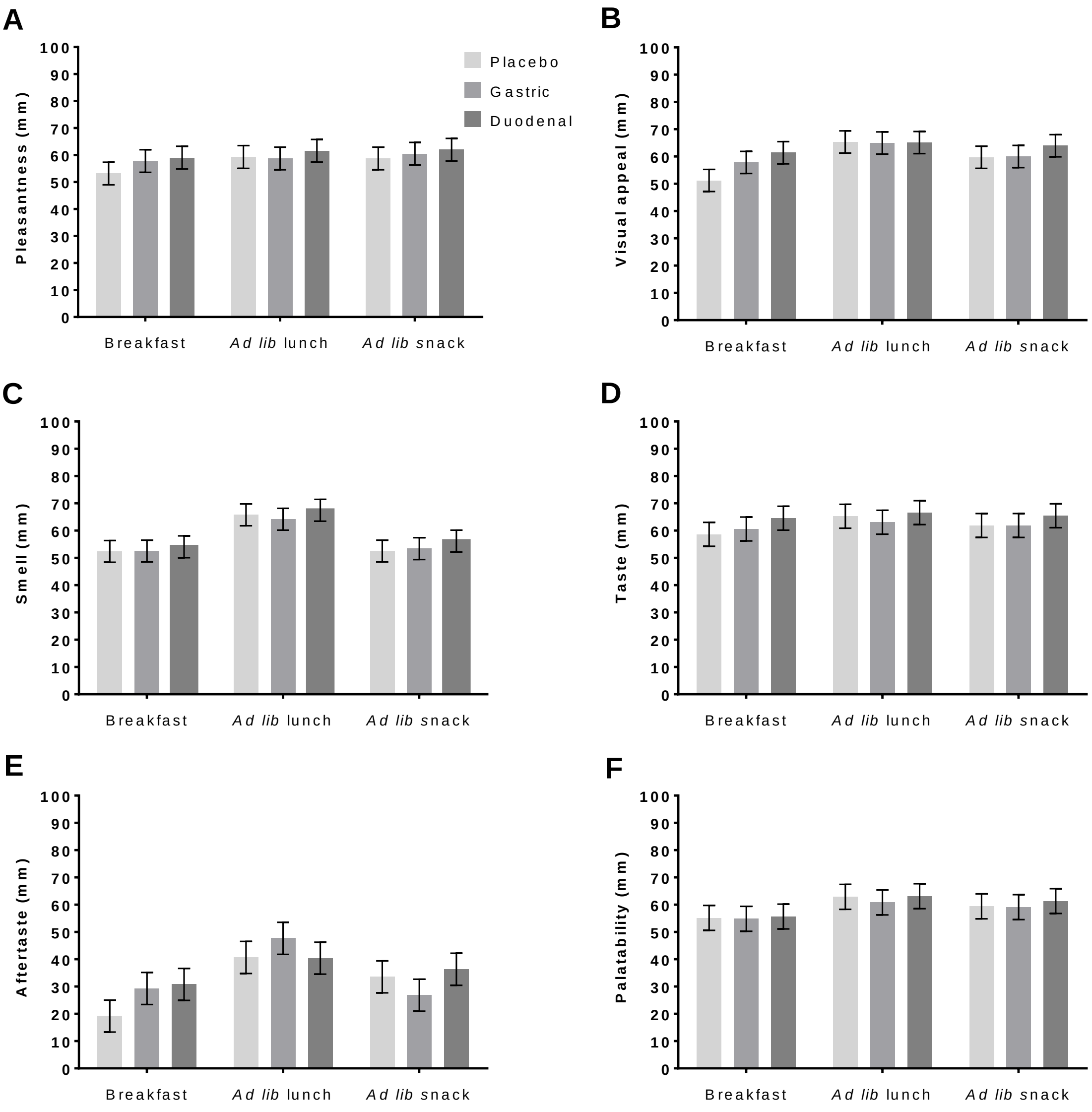


**Supplemental Figure 4** VAS ratings of (A) pleasantness, (B) visual appeal, (C) smell, (D) taste, (E) aftertaste, and (F) overall palatability obtained after the 2 MJ fixed energy breakfast (Breakfast) and the *ad libitum* lunch (*Ad lib* lunch) or snack (*Ad lib* snack). Treatments comprised administration of a vehicle control (Placebo) or Amarasate® targeted to either the small intestine (Duodenal) or stomach (Gastric) using delayed-release or standard capsules, respectively. Analysis was conducted using the mixed procedure (SAS 9.4) with treatment, visit number and meal (breakfast, *ad lib* lunch and *ad lib* snack) as factors. No significant effect of treatment was seen for any measure. Values are means ± SEM; n = 19. *Ad lib*, *ad libitum.*

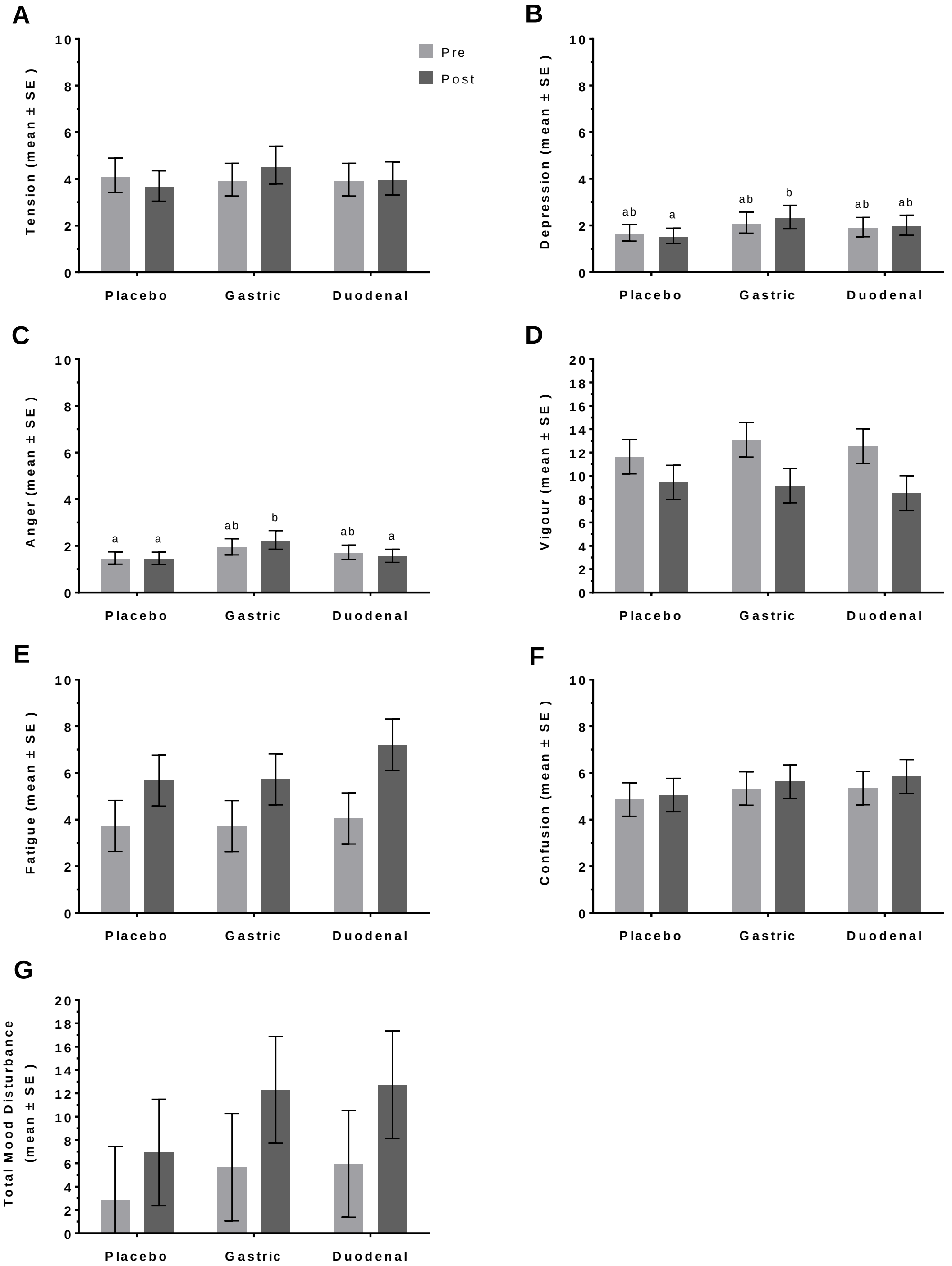


**Supplemental Figure 5** Effect of treatment on changes in mood state assessed using the Profile of Mood States (POMS) questionnaire subscales (A) Tension-Anxiety, (B) Depression-Dejection, (C) Anger-Hostility, (D) Vigor-Activity, (E) Fatigue-Inertia, (F) Confusion-Bewilderment, and the computed (G) total mood disturbance. Participants were asked to rate "how are you feeling right now?". The questionnaire was administered 1 h before (Pre, T = -60 min) receiving the treatment and repeated at T = 270 min (Post). Treatments comprised administration of a vehicle control (Placebo) or Amarasate® targeted to either the small intestine (Duodenal) or stomach (Gastric) using delayed-release or standard capsules, respectively. Analysis was conducted using the mixed procedure (SAS 9.4) with treatment, visit number and time (pre/post) as factors. A significant effect of treatment was observed for B and C ($p < 0.05$) only. Letters denote significantly ($p < 0.05$) different means using Fisher's LSD *post hoc* pairwise comparisons. Values are means ± SEM; n = 19.
